# Prevalence, Patterns and Predictors of physical activity in Urban Population of Bhubaneswar smart city, India

**DOI:** 10.1101/2020.11.28.20240358

**Authors:** Satyajit Mohanty, Jyotiranjan Sahoo, Epari Venkatarao, G Ganesh Shankar, Sandeep Kumar Panigrahi

**Author notes:** **Corresponding author:** Dr. Sandeep Kumar Panigrahi, Associate Professor, Community Medicine Department, IMS and SUM Hospital, Siksha O Anusandhan deemed to be University, Bhubaneswar, Odisha, India – 751003.

## Abstract

**Background:** Physical inactivity is a risk factor for mortality and morbidity. Physical activity and its predictors among urban population in this part of the country was unknown. Finding physical inactivity as a cause of current noncommunicable diseases (NCD) is difficult.

**Objectives:** To find out the prevalence, patterns and predictors of physical activity in urban population, and investigate its causal relationship with NCD.

**Materials and methods:** It was a cross-sectional study using cluster random sampling. Sample size was 1203. Socio-demographic, health profile, physical activity levels, and stage of change for physical activity behaviour were collected. was used for analysis. Logistic regression and marginal structural model analysis (by IPTW) were done using IBM SPSS 20.0.Statistical significance were tested at p=0.05.

**Results:** 1221 subjects participated. Mean age was 35.25 years. 71.9% were physically inactive, 15.9% practised ‘yogasana’. General caste, presence of NCD, being in a static stage of change and a yogassana practitioner influenced physical activity positively. Physical inactivity had 1.54 times higher odds for NCD and was statistically significant.

**Conclusion:** Prevalence of physical activity was low. Physical inactivity was a causative factor for NCD.

## Introduction

Physical activity (PA) is defined as any bodily movement produced by skeletal muscles that require energy expenditure, which includes structured activity programs such as exercises, and activities undertaken while working, playing, and carrying out household works [1]. Regular PA has been found to improve physiological functioning, quality of life, social and work participation [2]. On the other hand unhealthy dietary habits, physical inactivity and genetic predisposition can result in worsening of several chronic conditions including type 2 diabetes mellitus, and cognitive disorders [3,4]. Insufficient physical activity is considered as one of the leading risk factors for global mortality and increases the disability-adjusted life years (DALYs) [5]. Physical inactivity is therefore considered a global public health problem [6]. Considering the overall benefits of PA, the feasibility to engage and low cost, the WHO has recommended a cumulative engagement of minimum 150 minutes per week of moderate physical activity [7]. Maintaining physical activity throughout life is considered an important public health objective [2,8].

Despite the documented benefits and importance, 23% of males and 32% females above 18 years of age are physically inactive around the world [7] and there is a global trend towards engaging in sedentary behaviours [9]. Compared to global estimates, Indians are found to be sedentary and less physically active. A multi-centre study conducted in four regions of India and involving 14,227 participants concluded that large percentage of people in India are inactive and less than 10% population engages in recreational PA [10]. Another work reported that the exercise intensities undertaken do not meet the global recommended intensities, even in those who undertake leisure time PA [11].

Apart from traditional PAs, there is a necessity to evaluate the prevalence of people who follow other complementary and alternative therapies such as yoga for engaging in PA, as it promotes holistic health. Though yoga may not be plainly compared to physical exercises, yoga interventions are equally effective to exercises in improving positive outcomes [12–14]. This data on the acceptability of Yoga as a tool may be important to policy makers to establish yoga wellness centers and provide yoga resources to the general public. Recognizing the importance of PA, the Government of India has launched the *‘Fit India movement ‘* [15] and the *‘Khelo India’ programme* [16] to promote fitness as a way to create a healthy society and strong country.

Why few people are physically active and others are inactive is an intriguing question. A number of theoretical models and numerous studies from western countries attempted to look into this aspect [17–19]. Past exercise behaviours, perceived self-efficacy, social support, self-confidence, access to facilities, physical environment, gender and socio-economic status are identified as some factors that can influence PA behaviour [18]. Lower levels of motivation, limited free time, fear of falling, cost, transportation, pain, and lack of enjoyment are seen as barriers to participating in regular PA [20–22]. A report from Kenya found associations between low levels of PA and the demographic and social changes that prevail there [23]. However, no previous work has specifically looked into the PA patterns and the factors that predict PA among the urban population in Odisha state of India. This may be deemed important as the factors that predict PA patterns may be different across cultures and data on PA are important to formulate action plans for the promotion of PA for a region. The objective of this study was therefore to evaluate the PA patterns and predict the factors that influence the PA pattern in an urban setting in Bhubaneswar city of Odisha, India.

NCDs account for 61% of total deaths in India and are attributed to lifestyle-related risk factors such as tobacco use, physical inactivity, alcohol consumption, and obesity [24,25]. A 2010 cross sectional study identified tobacco and alcohol use, low intake of fruit and vegetables, and underweight as risk factors amongst the lower socioeconomic positions in rural Indians, while obesity, dyslipidaemia, and diabetes in men and hypertension in women as the more prevalent risk factors for NCDS for those in higher socioeconomic positions [26]. Though promoting better urban design to increase physical activity and reducing sedentary lifestyles in Urban India has been put forward as a strategy to reduce the risk of NCDs [27], we could not identify literature that have examined the relationship between physical activity and NCDs in India either as an independent risk factor or when controlled for other variables. Therefore, we proposed to use inverse probability of treatment weighting (IPTW) method to investigate the relationship between physical activity and prevalence of NCDs in a large urban based study.

### Objectives

1. *To find out the prevalence and patterns of physical activity in urban population.*
2. *To investigate the relationship of non-communicable disease with physical inactivity.*

## Methods

The study participants were recruited from Bhubaneswar, the capital city of State of Odisha, India. This cross-sectional survey was conducted from February to August 2019 involving adults of both sexes aged 18 years and above. Bhubaneshwar has a population of 8,85,363 residing in 1,97,661 households spread over 67 administrative wards and about 18.5% of the population reside in the slums[28,29].

The sample size was calculated based on previous estimates of the prevalence of physical activity (45.6%) in an urban setting[30]. Using a Z value of 1.96 at 95% confidence, absolute allowable error at 5% with an effect size of 3, the sample size was calculated to be 1203 to achieve 80% power.

30 administrative wards were randomly selected and the households were selected using a systematic random sampling method. Door-to-door visits were conducted between 2 PM to 7 PM to collect data by 2 enumerators. If a household refused to participate, the data was collected from the immediate next household. Only one response per household was documented. As the objective of study was to document the prevailing PA patterns, individuals with physical and mental disabilities who could not undergo regular PA otherwise were excluded from the study. Approval was obtained from the institute ethics committee prior to study commencement (DMR/IMS-SH/SOA/170066) and verbal informed consent was obtained from all participants in the local language. Though participants in the study were public, yet it was not appropriate or possible to involve patients or the public in the design, or conduct, or reporting, or dissemination plans of our research.

All the data were collected by the first author while another author oversaw the procedure. A pre-tested, semi-structured questionnaire was used to collect data on socio-demographic parameters related to age, religion, caste, marital status, education, occupation, family size, and monthly family income. Respondents were asked if they were suffering from any chronic diseases such as hypertension, diabetes, cardiovascular disease, asthma, and other chronic respiratory problems. Questions pertaining to addiction to alcohol, and smoking etc were collected along with factors that promoted PA. The behavioural aspects related to stages of change to PA promotion was evaluated using the Prochaska and DiClemente’s model [31]. The International physical activity questionnaire (IPAQ)-short from was used to assess the PA level of the participants. The IPAQ-short form is a 7-day physical activity recall questionnaire which has seven questions, related to frequency, duration and intensity of PA[32]. The IPAQ-short form is found to be valid and reliable [33–35]. A question on the current practice of yoga was asked. The average time taken to complete the questionnaire was 15 minutes.

### Classifications used

The socioeconomic status (SES) of the participants was determined using modified BG Prasad SES scale (2018) based on per capita monthly income. The SES was classified into five groups, namely lower, upper-lower, middle, upper-middle and upper class[36]. Chronic diseases were classified as per the International statistical classification of diseases and related health problems-10th revision (ICD-10) [37]. The state of change with respect to PA/exercise was grouped into either static (Pre-contemplation and Relapse) or dynamic (Contemplation, Decision, Action, and Maintenance) stages [38]. Based on the IPAQ scores, PA levels were classified as low, moderate and high. Participants who were classified under moderate PA and high PA were considered to be ‘physically active’ for the purpose of this study. Respondents classified as having low PA was considered as ‘physically inactive’.

### Statistical analysis

Statistical analyses were performed using the SPSS software version 20 (IBM SPSS). Categorical variables were expressed in terms of frequency and percentages. Estimates were expressed as mean ± standard deviation or proportions. Association between two categorical variables was tested using the Chi-squared test. Presence of gender bias was tested during analysis. Logistic regression analyses were used to compute adjusted odds ratios for each variable. *P*-values of less than 0.05 were considered significant.

The causality of chronic disease by ‘physically inactive’ was tested through inverse probability of treatment weighting (IPTW) using marginal structural model (MSM) using a Generalized Estimating Equation (GEE). The covariates used to generate the propensity score was the same variables found to be associated with physically activity status in univariate analysis (with p value of less than 0.20). First the weighted predicted values (inverse propensity score) were calculated for physical activity categories which was later normalized through robust estimator for the covariance matrix applying generalized estimating equations for predicting association of insufficiently physically active to that of chronic diseases.

## Results

A total of 1221 study participants were interviewed from the selected thirty clusters. After data cleaning, a total 1125 cases were included for analysis. Mean age of the study participants was 35.25 (± SD 10.72) years. Males and females gender representation in the group was not significantly different (p=0.350). The average distance of the nearest public exercise facility from home was 496.81 (± SD 238.73) meters with a maximum distance of 1500 meters. Majority (87.4%) of the study population was in the dynamic stage of exercise behaviour change according to the Prochaska and DiClemente model. A total of 809 (71.9%) of the respondents were classified as physically inactive. Rest 316 (28.1%) of the respondents were found to be physically active among and them only 11 (1%) were involved in high PA. 179 (15.9%) respondents practised ‘yogasana’ (Table 1). Among the practitioners of yoga, 68 (32.8%) met recommended standards of PA.

**Table 1:**
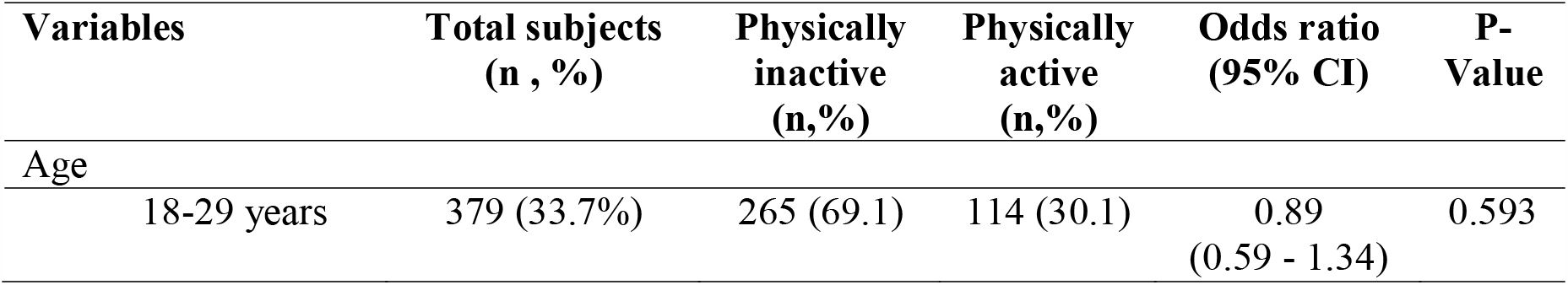
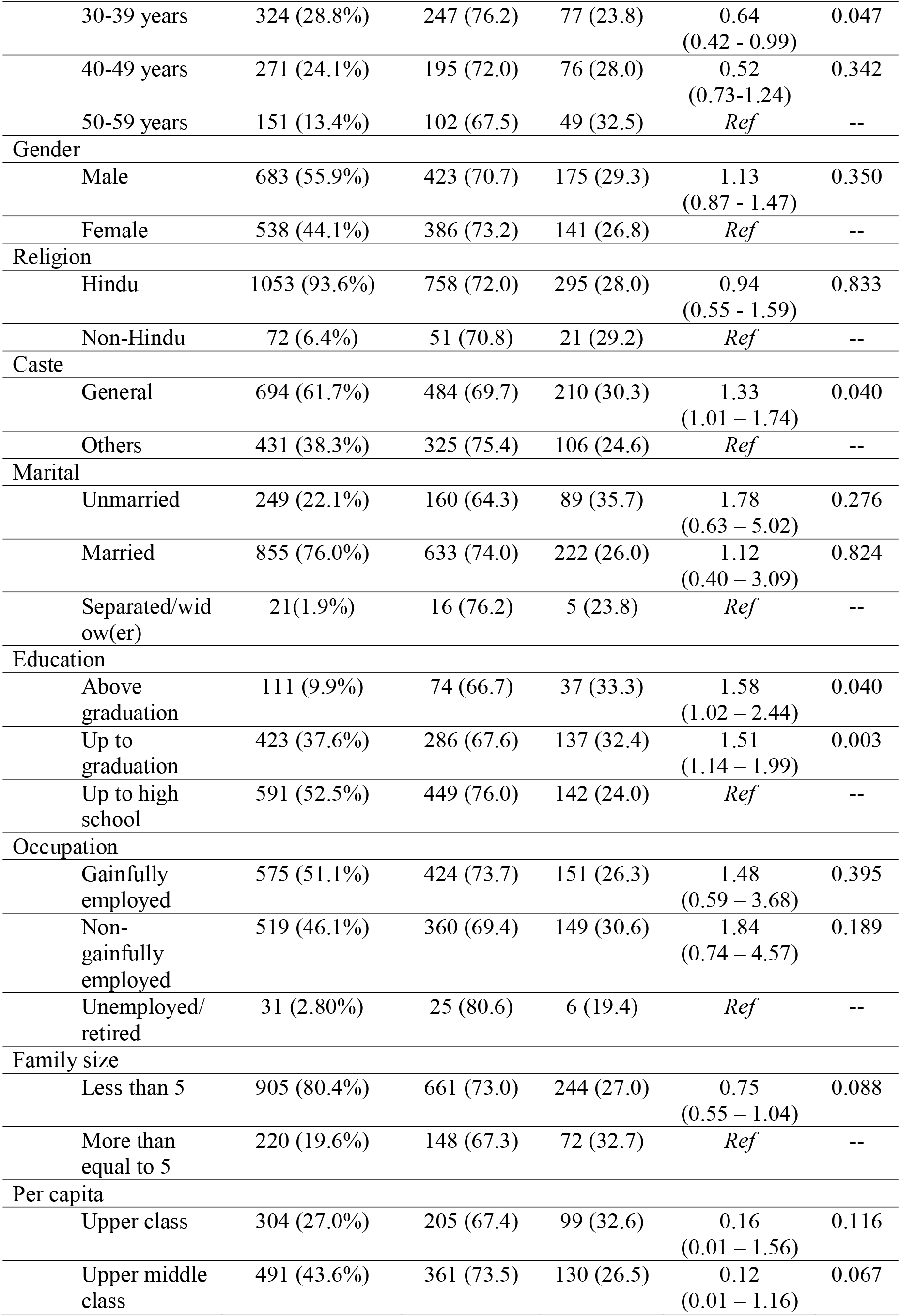
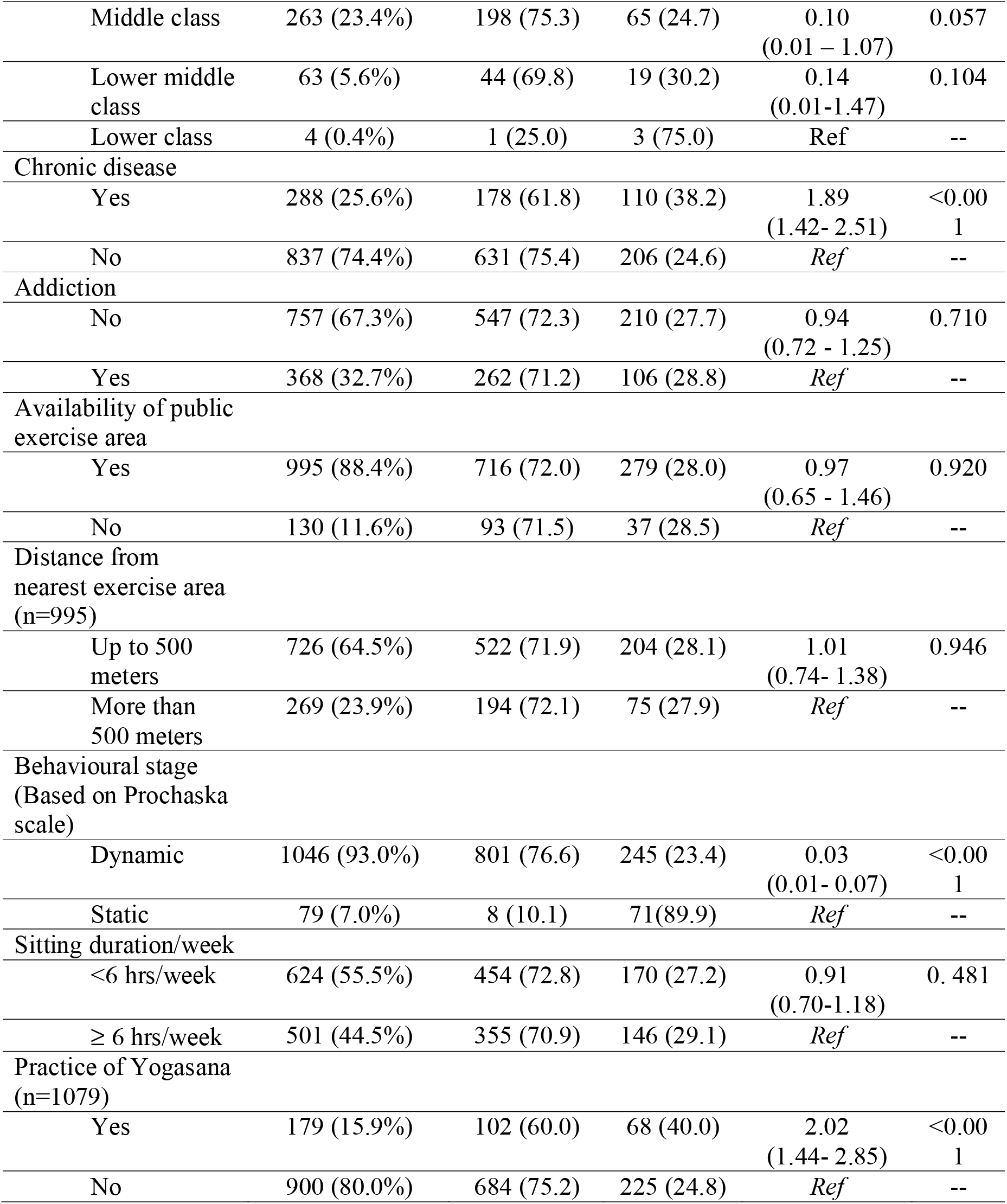
Characteristics of the study population (n=1125)

| Variables | Total subjects<br>(n , %) | Physically<br>inactive<br>(n,%) | Physically<br>active<br>(n,%) | Odds ratio<br>(95% CI) | P-Value |
| --- | --- | --- | --- | --- | --- |
| Age |  |  |  |  |  |
| 18-29 years | 379 (33.7%) | 265 (69.1) | 114 (30.1) | 0.89<br>(0.59 - 1.34) | 0.593 |
| 30-39 years | 324 (28.8%) | 247 (76.2) | 77 (23.8) | 0.64<br>(0.42 - 0.99) | 0.047 |
| 40-49 years | 271 (24.1%) | 195 (72.0) | 76 (28.0) | 0.52<br>(0.73-1.24) | 0.342 |
| 50-59 years | 151 (13.4%) | 102 (67.5) | 49 (32.5) | *Ref* | -- |
| **Gender** |  |  |  |  |  |
| Male | 683 (55.9%) | 423 (70.7) | 175 (29.3) | 1.13<br>(0.87 - 1.47) | 0.350 |
| Female | 538 (44.1%) | 386 (73.2) | 141 (26.8) | *Ref* | -- |
| **Religion** |  |  |  |  |  |
| Hindu | 1053 (93.6%) | 758 (72.0) | 295 (28.0) | 0.94<br>(0.55 - 1.59) | 0.833 |
| Non-Hindu | 72 (6.4%) | 51 (70.8) | 21 (29.2) | *Ref* | -- |
| **Caste** |  |  |  |  |  |
| General | 694 (61.7%) | 484 (69.7) | 210 (30.3) | 1.33<br>(1.01 – 1.74) | 0.040 |
| Others | 431 (38.3%) | 325 (75.4) | 106 (24.6) | *Ref* | -- |
| **Marital** |  |  |  |  |  |
| Unmarried | 249 (22.1%) | 160 (64.3) | 89 (35.7) | 1.78<br>(0.63 – 5.02) | 0.276 |
| Married | 855 (76.0%) | 633 (74.0) | 222 (26.0) | 1.12<br>(0.40 – 3.09) | 0.824 |
| Separated/widow(er) | 21(1.9%) | 16 (76.2) | 5 (23.8) | *Ref* | -- |
| **Education** |  |  |  |  |  |
| Above graduation | 111 (9.9%) | 74 (66.7) | 37 (33.3) | 1.58<br>(1.02 – 2.44) | 0.040 |
| Up to graduation | 423 (37.6%) | 286 (67.6) | 137 (32.4) | 1.51<br>(1.14 – 1.99) | 0.003 |
| Up to high school | 591 (52.5%) | 449 (76.0) | 142 (24.0) | *Ref* | -- |
| **Occupation** |  |  |  |  |  |
| Gainfully employed | 575 (51.1%) | 424 (73.7) | 151 (26.3) | 1.48<br>(0.59 – 3.68) | 0.395 |
| Non-gainfully employed | 519 (46.1%) | 360 (69.4) | 149 (30.6) | 1.84<br>(0.74 – 4.57) | 0.189 |
| Unemployed/retired | 31 (2.80%) | 25 (80.6) | 6 (19.4) | *Ref* | -- |
| **Family size** |  |  |  |  |  |
| Less than 5 | 905 (80.4%) | 661 (73.0) | 244 (27.0) | 0.75<br>(0.55 – 1.04) | 0.088 |
| More than equal to 5 | 220 (19.6%) | 148 (67.3) | 72 (32.7) | *Ref* | -- |
| **Per capita** |  |  |  |  |  |
| Upper class | 304 (27.0%) | 205 (67.4) | 99 (32.6) | 0.16<br>(0.01 – 1.56) | 0.116 |
| Upper middle class | 491 (43.6%) | 361 (73.5) | 130 (26.5) | 0.12<br>(0.01 – 1.16) | 0.067 |
| Middle class | 263 (23.4%) | 198 (75.3) | 65 (24.7) | 0.10<br>(0.01 – 1.07) | 0.057 |
| Lower middle class | 63 (5.6%) | 44 (69.8) | 19 (30.2) | 0.14<br>(0.01-1.47) | 0.104 |
| Lower class | 4 (0.4%) | 1 (25.0) | 3 (75.0) | Ref | -- |
| Chronic disease |  |  |  |  |  |
| Yes | 288 (25.6%) | 178 (61.8) | 110 (38.2) | 1.89<br>(1.42- 2.51) | <0.001 |
| No | 837 (74.4%) | 631 (75.4) | 206 (24.6) | Ref | -- |
| Addiction |  |  |  |  |  |
| No | 757 (67.3%) | 547 (72.3) | 210 (27.7) | 0.94<br>(0.72 - 1.25) | 0.710 |
| Yes | 368 (32.7%) | 262 (71.2) | 106 (28.8) | Ref | -- |
| Availability of public exercise area |  |  |  |  |  |
| Yes | 995 (88.4%) | 716 (72.0) | 279 (28.0) | 0.97<br>(0.65 - 1.46) | 0.920 |
| No | 130 (11.6%) | 93 (71.5) | 37 (28.5) | Ref | -- |
| Distance from nearest exercise area (n=995) |  |  |  |  |  |
| Up to 500 meters | 726 (64.5%) | 522 (71.9) | 204 (28.1) | 1.01<br>(0.74- 1.38) | 0.946 |
| More than 500 meters | 269 (23.9%) | 194 (72.1) | 75 (27.9) | Ref | -- |
| Behavioural stage (Based on Prochaska scale) |  |  |  |  |  |
| Dynamic | 1046 (93.0%) | 801 (76.6) | 245 (23.4) | 0.03<br>(0.01- 0.07) | <0.001 |
| Static | 79 (7.0%) | 8 (10.1) | 71(89.9) | Ref | -- |
| Sitting duration/week |  |  |  |  |  |
| <6 hrs/week | 624 (55.5%) | 454 (72.8) | 170 (27.2) | 0.91<br>(0.70-1.18) | 0.481 |
| ≥ 6 hrs/week | 501 (44.5%) | 355 (70.9) | 146 (29.1) | Ref | -- |
| Practice of Yogasana (n=1079) |  |  |  |  |  |
| Yes | 179 (15.9%) | 102 (60.0) | 68 (40.0) | 2.02<br>(1.44- 2.85) | <0.001 |
| No | 900 (80.0%) | 684 (75.2) | 225 (24.8) | Ref | -- |

Mean days/week spent in walking was 1.98 (95% CI: 1.80 – 1.16) with mean walk duration was 17.56 (95% CI: 16.76 – 18.37) mins/day. Mean days/week spend in ‘moderate intensity’ PA was 0.33 (95% CI: 0.25 – 0.41) with mean ‘moderate intensity’ PA duration (min) /day was 12.56 (95% CI: 11.88 – 13.25). Mean days/week spend in ‘high intensity’ PA was 0.19 (95% CI: 0.13 – 0.25) with mean ‘high intensity’ PA duration (min)/day was 11.44 (95% CI: 10.92 – 11.95).

### Univariate analysis

Caste, Educational attainment, Chronic disease, Stage of change and practice of yogasana are found to be associated with physical activity. Whereas, gender, religion, marital status, SES, employment status, family size, addiction, cumulative sitting duration, presence of neighbourhood public exercise facility or availability of neighbourhood exercise facility within 500 meters do not have any association with physical activity (Table 1).

### Multivariate analysis

After adjusting for confounding factors social caste, chronic disease status, practice of yogasana and stage of change were found to be associated with physical activity. Caste, chronic disease status, practice of yogasana were the positive predictors whereas stage of change was found to be a negative predictor of physical activity in our study. General caste had 1.43 times higher odds of being physically active compared to reserved which was statistically significant (95% CI: 1.03 – 1.99, P = 0.031). Respondents with chronic disease and practice of yogasana had higher odds of being physically active i.e. 2.08 (95% CI: 1.46 – 2.98) and 1.97 (95% CI: 1.35 – 2.89) respectively. Dynamic stage of change of exercise behaviour had lower odds of being physically active i.e. 0.02 (95% CI: 0.01 – 0.05). A physically active person in Bhubaneswar was more likely from general caste, having some form of chronic disease, in the static stage of change and a yogasana practitioner. Age, education, employment status, family size and SES did not exhibit any association with physical activity status (Table 2).

**Table 2.**
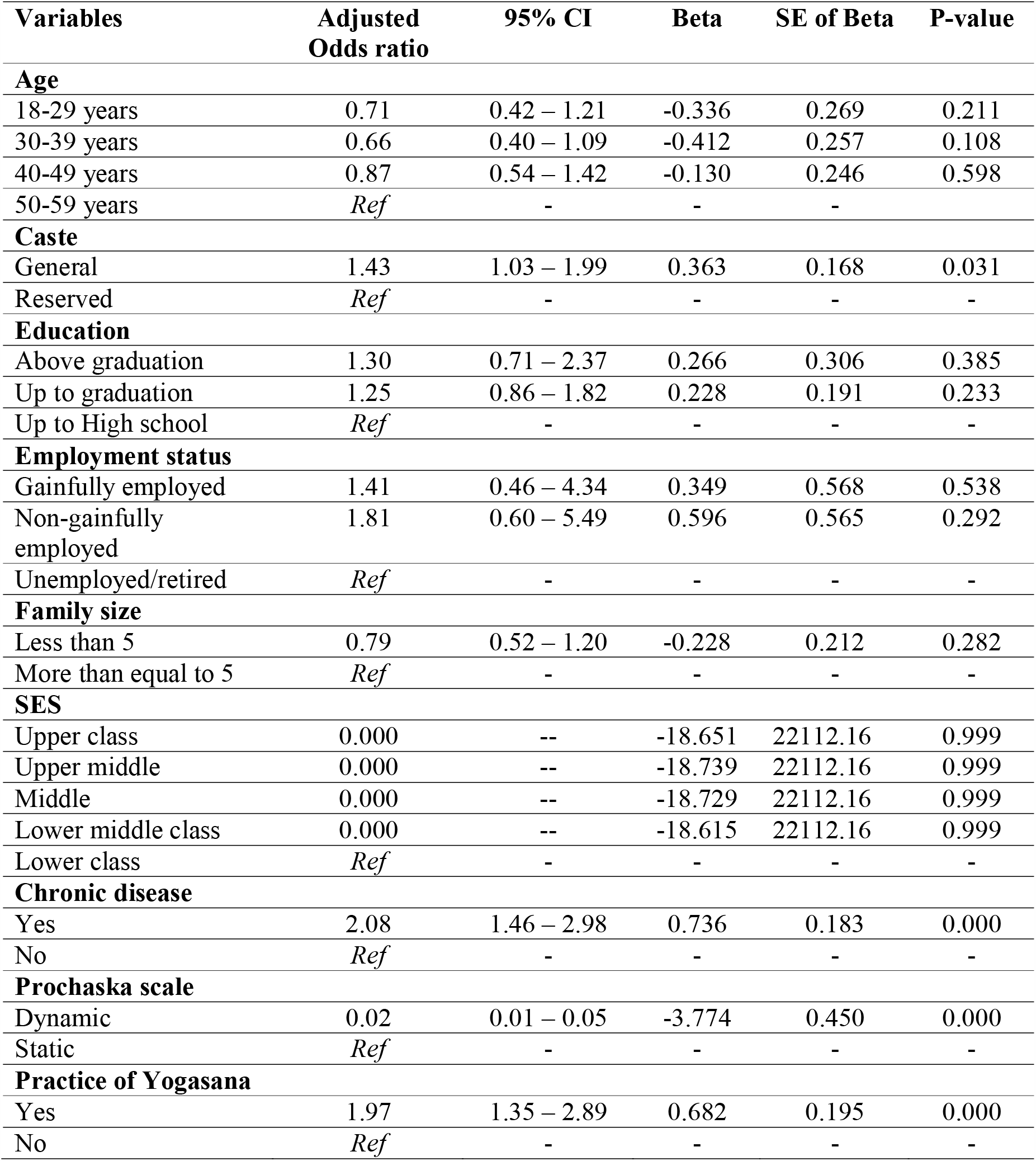
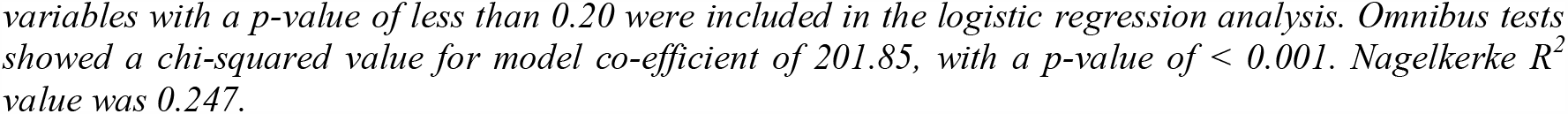
Binomial logistic regression analysis of different factors associated with physical activity.

| **Variables** | **Adjusted Odds ratio** | **95% CI** | **Beta** | **SE of Beta** | **P-value** |
| --- | --- | --- | --- | --- | --- |
| **Age** |  |  |  |  |  |
| 18-29 years | 0.71 | 0.42 – 1.21 | -0.336 | 0.269 | 0.211 |
| 30-39 years | 0.66 | 0.40 – 1.09 | -0.412 | 0.257 | 0.108 |
| 40-49 years | 0.87 | 0.54 – 1.42 | -0.130 | 0.246 | 0.598 |
| 50-59 years | *Ref* | - | - | - | - |
| **Caste** |  |  |  |  |  |
| General | 1.43 | 1.03 – 1.99 | 0.363 | 0.168 | 0.031 |
| Reserved | *Ref* | - | - | - | - |
| **Education** |  |  |  |  |  |
| Above graduation | 1.30 | 0.71 – 2.37 | 0.266 | 0.306 | 0.385 |
| Up to graduation | 1.25 | 0.86 – 1.82 | 0.228 | 0.191 | 0.233 |
| Up to High school | *Ref* | - | - | - | - |
| **Employment status** |  |  |  |  |  |
| Gainfully employed | 1.41 | 0.46 – 4.34 | 0.349 | 0.568 | 0.538 |
| Non-gainfully employed | 1.81 | 0.60 – 5.49 | 0.596 | 0.565 | 0.292 |
| Unemployed/retired | *Ref* | - | - | - | - |
| **Family size** |  |  |  |  |  |
| Less than 5 | 0.79 | 0.52 – 1.20 | -0.228 | 0.212 | 0.282 |
| More than equal to 5 | *Ref* | - | - | - | - |
| **SES** |  |  |  |  |  |
| Upper class | 0.000 | -- | -18.651 | 22112.16 | 0.999 |
| Upper middle | 0.000 | -- | -18.739 | 22112.16 | 0.999 |
| Middle | 0.000 | -- | -18.729 | 22112.16 | 0.999 |
| Lower middle class | 0.000 | -- | -18.615 | 22112.16 | 0.999 |
| Lower class | *Ref* | - | - | - | - |
| **Chronic disease** |  |  |  |  |  |
| Yes | 2.08 | 1.46 – 2.98 | 0.736 | 0.183 | 0.000 |
| No | *Ref* | - | - | - | - |
| **Prochaska scale** |  |  |  |  |  |
| Dynamic | 0.02 | 0.01 – 0.05 | -3.774 | 0.450 | 0.000 |
| Static | *Ref* | - | - | - | - |
| **Practice of Yogasana** |  |  |  |  |  |
| Yes | 1.97 | 1.35 – 2.89 | 0.682 | 0.195 | 0.000 |
| No | *Ref* | - | - | - | - |

variables with a *p*-value of less than 0.20 were included in the logistic regression analysis. Omnibus tests showed a chi-squared value for model co-efficient of 201.85, with a *p*-value of < 0.001. Nagelkerke *R*<sup>2</sup> value was 0.247.

### IPTW analysis

Physically inactive subjects have higher odds (adjusted OR = 1.54) of having chronic disease (Table 3). Prior established determinants of the chronic disease are considered for our analysis i.e. age, gender, behaviour, marital status, education, occupation, neighbourhood characteristics (Presence of common exercise area), addiction, sitting time. Inverse probability of treatment weighing method (IPTW) and later generalized estimating equations (GEE) was used to explore the causal association between physical inactivity and chronic diseases based on above mentioned factors.

**Table 3:** Association of physical inactivity and chronic diseases (n=1125)

| **Variables** | **Adjusted Odds ratio** | **95% CI** | **Beta** | **SE of Beta** | **P-value** |
| --- | --- | --- | --- | --- | --- |
| Physically inactive | 1.54 | 1.004 – 2.382 | 0.436 | 0.220 | 0.048 |

## Discussion

The study had a uniform representation of the study population of the urban city. There was no inclusion bias for gender as was seen during analysis (p>0.05). The main results of the study show that the over-all level of PA undertaken by the general public in Bhubanewar is low. A mere 28.1%, (316/1125) participants were found to be physically active as measured by the IPAQ-short form. These scores are low compared to a work that reported 38.3% of the population to indulge in sufficient PA in India (29). Another international work that assessed PA from different countries found that only 9.3% men and 15.2% women are physically inactive (28) in India. However, another study conducted in an urban city in South India reported 49.7% prevalence of physical inactivity in a sample of 286 adults as measured by the WHO standard Global Physical Activity Questionnaire [41]. On the contrary, the results from a market intelligence agency showed that 64% Indians in a sample of 3000 adults aged 18 and above do not exercise [42]. This inconsistency in reporting PA prevalence patterns in India are perplexing. We hypothesize this to be attributed to the study population, location of study (urban/ rural), type of questionnaire used to document PA or there may be rapidly growing physical inactivity in the Urban Odisha.

Another alarming result that we found from our study was the 25.6% prevalence rate of participants suffering from at least a chronic disease. A recent work estimated the overall prevalence of diabetes, hypertension, and obesity in India as 2.9%, 14.4% and 9.7%, respectively [43]. The results of our study showed that persons who suffer from chronic disorders are at increased odds of engaging in a regular PA program. This either we hypothesize to the prescription or advice by HCPs to participate in PA or the participants might have started doing exercises understanding the benefits of PA. This is in contrary to previous results which found that the activity limitations increases with the number of chronic conditions and reduces the amount of PA undertaken in people with chronic disorders[44,45].

368 (32.7%) of the participants self-reported to be addicted to illicit substances that are identified as risk factors that contribute to non-communicable diseases. This number (32.7%) is higher than the combined risk factors (alcohol use and tobacco use) provided in the 2014 report on global status of non-communicable diseases [46]. Studies have shown mixed-results regarding the association between substance abuse and PA. One study reported positive association of PA with current and future alcohol use but negative association with tobacco and other drug use [47]. Literature consistently reports that PA is inversely associated with tobacco use in adults and physically active adults are more likely to be moderate drinkers [48,49].

Our study results show that while education can influence PA pattern individually, the role of education seems to be silent when controlled for other sociodemographic factors. We hypothesize this to the role of perceived self-control executed by education on behavioural change [50]. Persons with low perceived control are found to lack confidence, and neither do not work for positive intended outcomes nor work towards elimination of undesired outcomes [51]. The study limitations such as having more than 50% study participants who were educated till graduation level, PA was self-reported and was not reporting any long-term status change, the period of data collection was immediately before COVID-pandemic with not so good national and global economic conditions might have played a role in the study period between predictors.

The results also showed that being in the dynamic stage of Prochaska stage of change was found to negatively predict PA. This scale describes how the attitude changes based on the duration of behavior change [51]. The reasons for the negative predictions are beyond the scope of study. However, past works have shown that such changes to be more effective in participants who are already assigned to an intervention, or in those who with pre-existing health disorders who have been assigned to an intervention [52–55].

The results that physically inactive participants have higher odds of having chronic disease are not surprising. PA is touted as a primary preventive measure from chronic disorders and fosters wellness in general [56–58]. PA also prevents the progression of symptomatic diseases and delay its progression to disability, or death [58,59].

Our data also showed a reduction in the duration and intensity of walking as measured by our study participants. The over-all walking percentage by humans have reduced by 50-70% and a mean cadence of 7473 steps has been reported from 26 studies conducted between 1966-2007 [60,61]. Normative data indicates that normal adults walk between 4,000 and 18,000 steps/day; and 10,000 steps/day are considered as reasonable cadence per day. Literature suggests that efforts have to be initiated to increase the walking to a minimum of 2,000-2,500 steps/day [62].

15.9% of our study respondents reported practicing yoga. Our figures are way higher than the 5.3% and 11.8% reported by a previous work in the eastern zone and pan-India respectively [62].The increased awareness could be attributed to the roots of yoga’s origins that can be traced to India and the national promotion towards celebration of International Day of Yoga and follow yoga as a daily part of life. The study results showed that those who practised yoga had 2.02 times higher odds of being physically active. This may be attributed to the role of yoga in increasing self-efficacy and intrinsic motivation, which may encourage persons to engage in PA [63]. Though it may be argued that subtle differences exist between physical exercises and the physical components of yoga practices, the results of our study shows yoga can be an effective and feasible alternative to PA.

Amongst other factors studied, multivariate analysis indicated that general caste had 1.43 times higher odds being physically active, when controlled for confounding factors. Social caste is a marker of socio-economic status [64] in India. Populations other than the general caste category fall into reserved caste and they represent the underserved population. Past work has reported that underserved populations were less likely to participate in sufficient moderate to vigorous PA [65].

There are some limitations to this study. For assessment of physical activity, IPAQ-short form was used, which is a self-reporting measure. The risks of recall bias leading to over- or under-reporting of PA cannot be ruled out and future works may consider using objective measures of PA, such as pedometers or accelerometers. Further, IPAQ is criticized for overestimating PA prevalence in population surveys [66]. Future studies may evaluate the associations between PA prevalence measured by IPAQ, GPAQ and objective measures. Though a representative population has been sampled, the results may be generalized to other urban places but not relevant to rural areas. Considering the differences in PA prevalence between the results of this study and other previous works, efforts should be made to study the national estimate using a common protocol.

## Conclusion

Prevalence of physically activity was found to be 28.1% out of which only 1% were involved in high level of Physical activity. 15.9% practised ‘yogasana’, with 32.8% of them meeting recommended standards of physical activity. General caste, already having a chronic disease, being in the static stage of change and a yogassana practitioner were all factors which made more likely for a person to be physically active. Physically inactive individuals were also found to have 1.54 times higher odds of having chronic disease than those who are physically active.

## Source of funding

None

## Data Availability

Data will be made available on request.

## Conflicts of interest

None

## Notes

### Competing Interest Statement

The authors have declared no competing interest.

### Author Declarations

Institute Ethics Committee of IMS and SUM Hospital vide letter number DMR/IMS-SH/SOA/170066

### Summary of Updates

Statements to include how selection bias was taken care of has been updated. Also statements related to use of public to design or conduct the research has been included. A statement on generalizability has been made more clear.

